# Applying the FAIR principles to data in a hospital: challenges and opportunities in a pandemic

**DOI:** 10.1101/2021.08.13.21262023

**Authors:** Núria Queralt-Rosinach, Rajaram Kaliyaperumal, César H. Bernabé, Qinqin Long, Simone A. Joosten, Henk Jan van der Wijk, Erik L.A. Flikkenschild, Kees Burger, Annika Jacobsen, Barend Mons, Marco Roos, BEAT-COVID Group, COVID-19 LUMC Group

**Affiliations:** Department of Human Genetics, Leiden University Medical Center,Leiden, The Netherlands; Department of Infectious Diseases, Leiden University Medical Center,Leiden, The Netherlands; Department of Biomedical Data Sciences, Leiden University Medical Center,Leiden, The Netherlands; Department of IT&DI, Leiden University Medical Center,Leiden, The Netherlands; GO FAIR Foundation, Leiden, The Netherlands; CODATA, Paris, France

**Keywords:** Patient Data, Ontologies, FAIR, Research Data Management, Hospital, Open Science

## Abstract

**Background:** The COVID-19 pandemic has challenged healthcare systems and research worldwide. Data is collected all over the world and needs to be integrated and made available to other researchers quickly. However, the various heterogeneous information systems that are used in hospitals can result in fragmentation of health data over multiple data ‘silos’ that are not interoperable for analysis. Consequently, clinical observations in hospitalised patients are not prepared to be reused efficiently and timely. There is a need to adapt the research data management in hospitals to make COVID-19 observational patient data machine actionable, i.e. more Findable, Accessible, Interoperable and Reusable (FAIR) for humans and machines. We therefore applied the FAIR principles in the hospital to make patient data more FAIR.

**Results:** In this paper, we present our FAIR approach to transform COVID-19 observational patient data collected in the hospital into machine actionable digital objects to answer medical doctors’ research questions. With this objective, we conducted a coordinated FAIRification among stakeholders based on ontological models for data and metadata, and a FAIR based architecture that complements the existing data management. We applied FAIR Data Points for metadata exposure, turning investigational parameters into a FAIR dataset. We demonstrated that this dataset is machine actionable by means of three different computational activities: federated query of patient data along open existing knowledge sources across the world through the Semantic Web, implementing Web APIs for data query interoperability, and building applications on top of these FAIR patient data for FAIR data analytics in the hospital.

**Conclusions:** Our work demonstrates that a FAIR research data management plan based on ontological models for data and metadata, open Science, Semantic Web technologies, and FAIR Data Points is providing data infrastructure in the hospital for machine actionable FAIR digital objects. This FAIR data is prepared to be reused for federated analysis, linkable to other FAIR data such as Linked Open Data, and reusable to develop software applications on top of them for hypothesis generation and knowledge discovery.

## Background

The COVID-19 pandemic has challenged healthcare and research data management systems worldwide to provide reusable patient data for rapid and efficient translational research. Clinical data, laboratory measurements, and various omics data such as transcriptomics and metabolomics, are routinely collected from hospitalized COVID-19 patients to inform medical doctors about patients’ health status and to support research on treatment options. Analysing data integrated from multiple sources in a hospital, complemented with data from other hospitals and public knowledge bases, can generate critical information about disease mechanisms to support diagnosis, prognosis and decisions on interventions. However, research and clinical data are often not prepared for instant secondary use involving multiple sources. This was already an obstacle for efficient clinical and biomedical research in general, but a pandemic of a poorly understood novel disease that overloads hospitals’ capacity has revealed the significance of this problem.

Integrative analysis is challenged by software systems used to collect these various types of data from patients in hospitals. Different formats may be used (e.g. CSV or JSON) and the semantics of data are often underspecified and captured in a proprietary syntax or by different standards (e.g. HL7 FHIR or OpenEHR). This can result in fragmentation over multiple ‘silos’ that are not sufficiently interoperable for instant computational analysis. Reuse and reproducibility are further hampered by missing or unstandardised provenance, such as the time and date at which data were collected (e.g. scans may be performed on a different day than blood measurements). Furthermore, to expand analysis beyond one hospital, information on consent and regulations that control data access, reuse, and sharing are often unclear and not easily assessable. Complete harmonization of access regulations between institutes and countries is not realistic, but analysis could still be efficient if access regulations were at least computationally assessable.

Ideally, hospital systems are set up with integrative, federated data analytics in mind. Global leaders in data science have posed that this can be achieved by applying agreed upon standards to make data globally findable, accessible, inter-operable, and reusable for humans and computers, also referred to by as ‘the FAIR principles’ [1]. Indeed, projects such as the GO FAIR Virus Outbreak Data Network (VODAN) [2], the ZonMW Covid program [3], the Trusted World of Corona (TWOC) [4], and ELIXIR Covid project [5] embrace FAIR principles as a key element of their COVID-19 data management strategy. A quintessential objective is turning data and data containers into machine actionable FAIR digital objects (FDO, in this paper defined as resources in a digital, machine understandable form; a formal framework for FDOs is under development, see [6, 7]). This will optimize the ability to integrate and visualise data from many sources, facilitate fine-grained data access regulation, and allow for decentralised and machine assisted analysis [8]. The latter is further enabled by the development of infrastructure that supports ‘data visiting’ [9, 10]. This is attractive for clinical data because (i) existing systems can be complemented with data visiting functions, thereby keeping their other functions in place, (ii) the output of an analysis is generally less privacy sensitive than the input. In Europe, the General Data Protection Regulation (GDPR) policy supports data visiting by requiring that access regulations for personal data are clearly defined [11].

Methods to facilitate the implementation of FAIR principles, or ‘FAIRification’, are currently being investigated in multiple projects and initiatives. We have previously published a generic workflow [12], as a basis for specialised variations such as for rare disease registries [13]. Related activities are the development of the FAIR cookbook in the FAIRplus project [14, 15], the three point framework for FAIRification of metadata by the VODAN GO FAIR network [16], and the organisation of a FAIRification steward team to support rare disease registries reach their FAIR goals [17]. The application of FAIR principles in hospitals is starting to be adopted in Europe as a key strategy for nationwide healthcare research data infrastructure [18, 19]. Cross connections through multinational collaborations, such as in ELIXIR and GO FAIR, and domain specific collaborations such as via globally operating patient organisations, could support convergence of FAIR implementation choices to further facilitate the adoption of FAIR principles and thereby efficient analysis across multiple hospitals in multiple countries.

At the Leiden University Medical Centre (LUMC), the implementation of FAIR principles for COVID-19 data is part of a multidisciplinary collaboration, coined ‘The BEAT-COVID project’. This collaboration was initiated in March 2020 to face the multiple analysis challenges of the COVID-19 pandemic. The LUMC is a tertiary care, teaching and research hospital in the Netherlands that encompasses clinical and research groups with expertise on immunology, biomedicine, data management and data science. The groups work together on collecting and sharing different types of patient data, analyses, findings, expertise, and novel solutions implemented in the hospital (e.g. see [20]). One of the challenges is to implement a FAIR Research Data Management plan (RDM) comprising FAIRification of priority resources and a FAIR based architecture that complements the existing data management systems in the hospital.

We hypothesise that the use of existing ontologies and ontological models will enable turning patient data into machine readable digital objects that are prepared for secondary use. Our objective is to develop ontological models that represent and link the data records and metadata of the datasets in the existing LUMC data management systems (Figure 1). In our ontology centred approach, data can stay in existing systems but are made accessible ‘in terms of’ the central data linking model to create a virtual warehouse. We reused existing ontological models such as the core ontological model for common data elements developed in the European Joint Programme on Rare Diseases (EJP RD) for patient registries [21], and the Data Catalogue Vocabulary (DCAT) for datasets [22]. The metadata is made accessible by a FAIR Data Point (FDP) instance [23]. FDPs ensure that BEAT-COVID resources can be found and used through querying machine readable metadata. It includes the pointers to access the content of the resource for analysis workflows, if access is permitted. By using ontologies, patient data in the hospital are virtually linked with other ontologically described data in the hospital, but also public Linked ‘Open’ Data (LOD). This can boost the potential for knowledge discovery and data+knowledge driven analytics. Interestingly, ontologies may also be used to describe data access restrictions [24, 25] to complement FAIR metadata with information that supports data safety and patient privacy.

**Figure 1.**
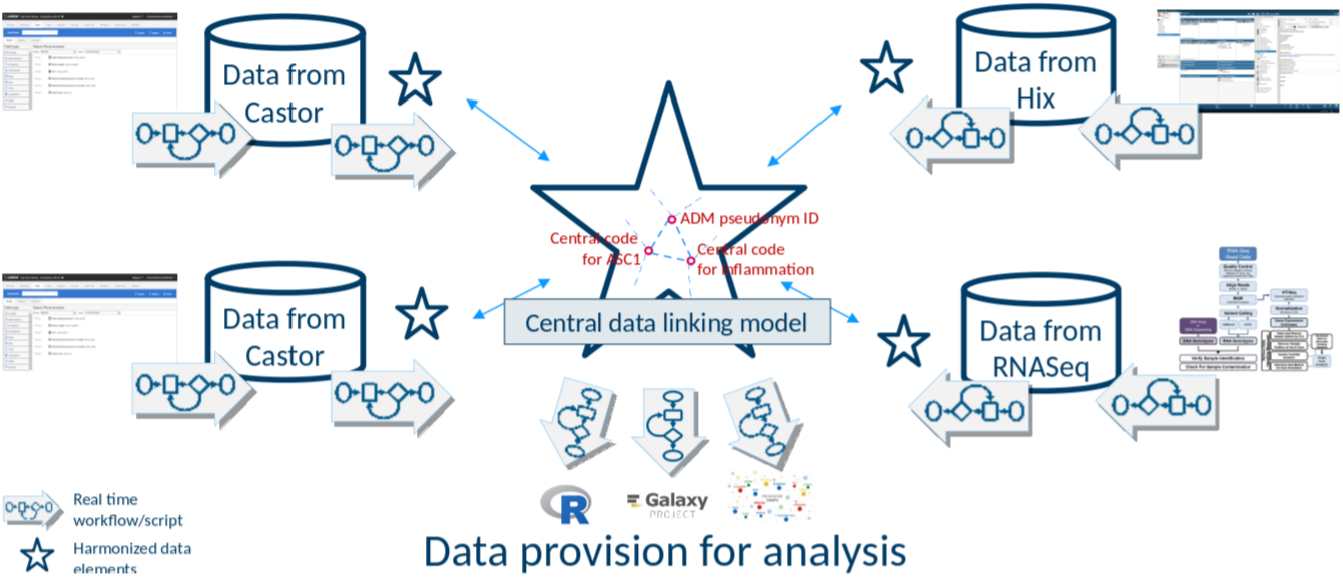
Illustration of the central concepts of the envisioned FAIR based architecture: the central star represents the data linking model for interoperability that the sources refer to (data and metadata), the small stars next to each source represent what is used of the central model to describe the source (thereby becoming ‘self-describing’), the arrows represent workflows or scripts: for the source systems to map or convert source data and metadata to the central data linking model, for retrieving data from across the sources through the central data linking model, and for analysis. FAIR Data Points provide access to the ‘ontologised’ metadata and data (not shown).

In this paper, we describe and implement our approach for FAIRification of COVID-19 observational patient data in an academic hospital. We selected cytokine measurements of hospitalised patients as our primary objective of FAIRification and development of the FAIR RDM. We synthesized an artificial dataset mimicking original laboratory data obtained from patient samples to study the data lifecycle without the risk of violating patient privacy. Our main result is the FAIRification in the hospital. We also show that our FAIRifcation approach is providing cytokine measurements as FDOs and is enabling applications on top of this FAIR patient data for analytics. Importantly, this work has been done in close collaboration with clinicians and data managers who are familiar with the existing hospital data systems and data lifecycles to establish best practices for making data FAIR in the hospital. We demonstrate that a FAIR RDM plan based on describing data and metadata by ontologies delivers an infrastructure that complements existing infrastructure with FDOs that are prepared for integrative and federated analysis. We show our first results and the solutions that are currently being developed as LUMC research data management procedures. We finally discuss what FAIRification entails in a ‘real world’ hospital situation involving different stakeholders and departments, and future challenges such as data access regulation in a FAIR ecosystem.

## Results

### FAIR status of patient data in existing systems

Our FAIR assessment of the cytokine data in existing systems revealed that while the structure and findability improved with each step of the data management lifecycle, no FAIR standards were applied to make the data and metadata globally understandable ‘for machines’, such as for automated computer processing (Table 1). The original data from the clinical laboratory that measured the cytokine levels was well structured, but not in a uniform, globally machine readable way. The data were further pre-processed manually and transferred to the Electronic Data Capture (EDC) software Castor [26]. Although this captured data electronically and in a uniform way, Castor does not apply FAIR standards. Data was subsequently transferred from Castor into the Opal data warehouse system [27], conform the standard workflow for preparing data for research at the LUMC. Opal is a generic system to bring datasets from different systems in the hospital into one warehouse, supporting transformations and annotation on the data level with a vocabulary chosen by the user. Opal provides researchers at the LUMC a central access point to research data that are syntactically machine readable. It offers APIs that bioinformaticians can incorporate in their workflows. Anonymised data of daily parameters from patient records was imported into Opal without including retrievable patient identifiers in the research environment of the hospital on almost real time.

**Table 1.**
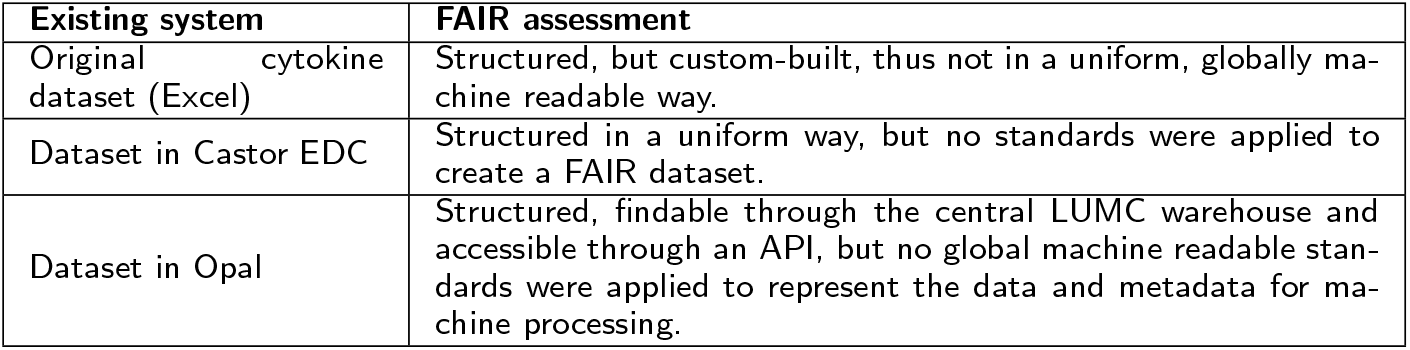
FAIR assessment of existing systems containing cytokine data.

| Existing system | FAIR assessment |
| --- | --- |
| Original cytokine dataset (Excel) | Structured, but custom-built, thus not in a uniform, globally machine readable way. |
| Dataset in Castor EDC | Structured in a uniform way, but no standards were applied to create a FAIR dataset. |
| Dataset in Opal | Structured, findable through the central LUMC warehouse and accessible through an API, but no global machine readable standards were applied to represent the data and metadata for machine processing. |

Opal’s native metadata tool Mica [28] provides annotation on the dataset level such as how, when, where, by whom, under what conditions data has been collected. This information is subsequently published in a Web portal. Therefore, Mica provides resource information that is human readable on the Web. This metadata is not also available in a machine readable form. Findability for machines can be improved by adding a machine readable ontological representation. Our automated FAIR assessment of a dataset described in Mica (see here) showed specifically which FAIR improvements could be made to make the metadata descriptions in Mica more machine actionable and standardized. Although Mica implements unique identifiers, these were not persistent in our case, and they were also not explicitly defined in the metadata. This creates challenges for data accessibility and reusability. Some systems, notably Opal [29], provide handles to integrate FAIR features, but we chose to first incorporate independent components to minimize requirements for other systems and thereby optimize reusability of the approach.

### Coordinated FAIRification

A coordinated FAIRification process with BEAT-COVID colleagues was set up to improve the machine readability, global interoperability, and findability of the COVID-19 data. We developed ontological models for data record in collaboration with data collectors, data managers, data analysts and medical doctors. Similarly, we developed machine actionable metadata to improve the findability, accessibility, and reusability of the datasets in collaboration with IT and database managers. Both tasks were performed in parallel and in a synergistic way to consistently support the entire data management lifecycle for data analysis, and they are ongoing for additional data types. While the BEAT-COVID project group was maintaining one-hour bi-weekly video calls for general update and logistic discussions, specific video calls were set up with the required experts and duration for the topic at hand. These regular and iterative meetings with all stakeholders were necessary to enable the development of optimal semantic modelling and computational standardization.

### Representing patient data as FAIR digital objects

Central to our approach to implementing FAIR principles ‘for machines’ is the composition of ontological models from existing commonly used ontologies. These models serve as reference for the data in the source systems, creating a larger ‘virtual’ data warehouse. In this section we present the ontological models and FAIR infrastructure that were set up to represent patient data as FDOs discoverable for analytics.

#### Ontological data model for interoperability of clinical measurements

To create a user centred research driven data infrastructure, we used the medical research questions as drivers for the data modelling. We first created a general concept model for the questions to extend with relevant clinical data, and mapped recurrent important terms mentioned by medical doctors into terms in Open Biological Biomedical Ontologies (OBO) ontologies [30] described in the Web Ontology Language (OWL) [31]. When we received the first actual data, cytokine measurements on samples collected from clinically admitted patients, we created an ontological model in Resource Description Framework (RDF) [32] for this data (see Figure 2). The cytokine model is based on the core semantic model that was developed in the EJP RD for common data elements in rare disease patient registries. This is a simple model that abstracts that every element in a patient registry is the outcome of a process, so that **process** becomes the core concept of the model [33]. We reused this model jointly with the quantitative trait semantic model [34] to capture clinical data measurements, where the ‘process of measurement’ is the core concept. Reusing these existing ontological models for observational data in the LUMC supports FAIR data. Not only does it allow interoperability with patient registries and quantitative traits, but also the common biomedical ontologies used allow data integration with external knowledge such as LOD.

**Figure 2.**
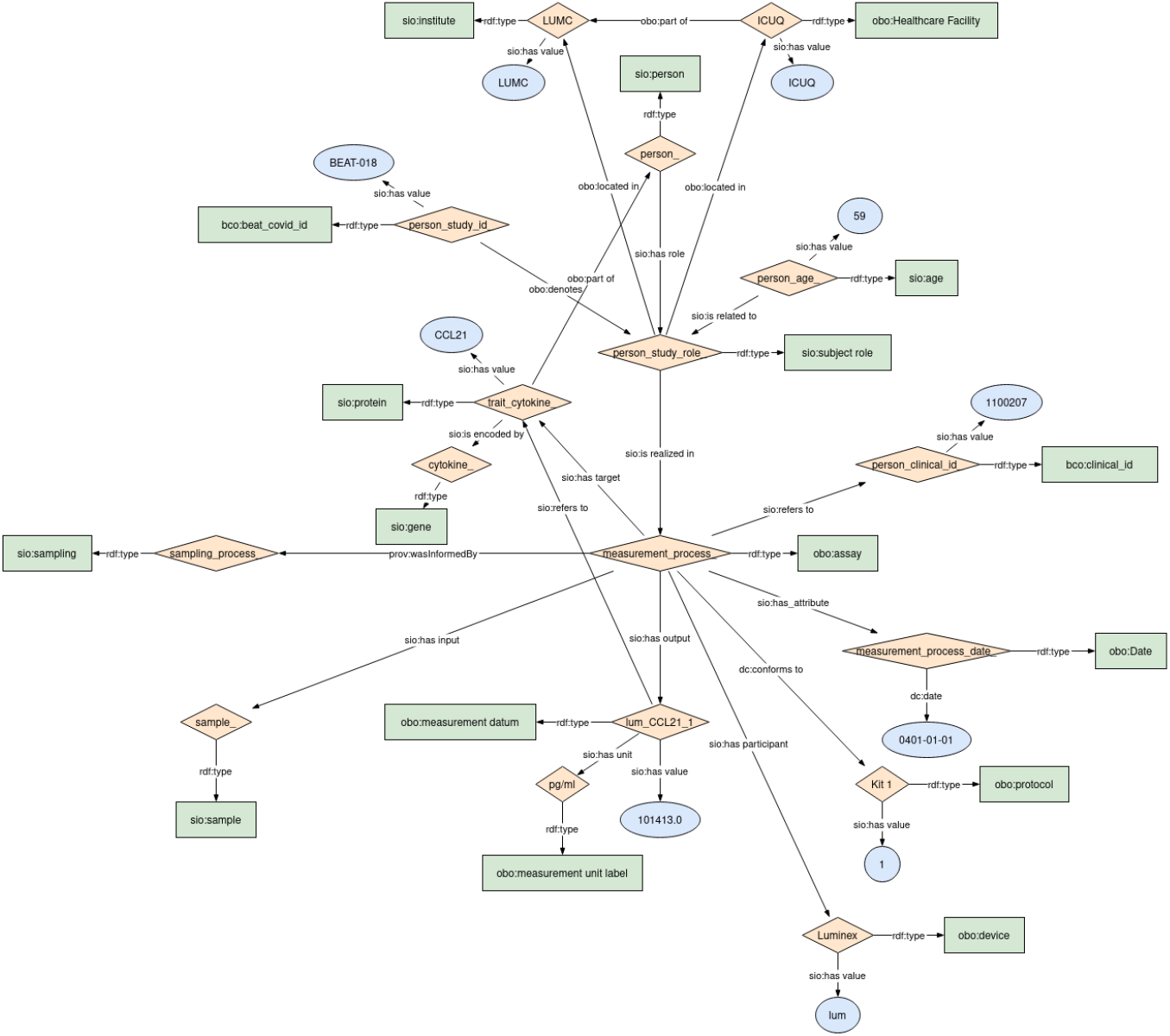
Ontological data model for the cytokine measurements patient dataset.

We also modelled a new semantic module for disease severity score phenotypes following the same EJP RD core model, see Figure 3. Apart from tracking the Apache IV Severity Score [35] and the SOFA Severity Score [36], medical doctors defined the Leiden Severity Score to obtain daily scores of disease severity for both COVID-19 patients admitted to the ward and ICU (Intensive Care Unit). All these scores are based on lab results and clinical data and reflect the actual disease severity of the patient on that day and are informative for doctors to make decisions about patient care management. The ontological linking data model, and its modules (lab measurements, biosamples and disease severity score), are publicly available on GitHub - data model.

**Figure 3.**
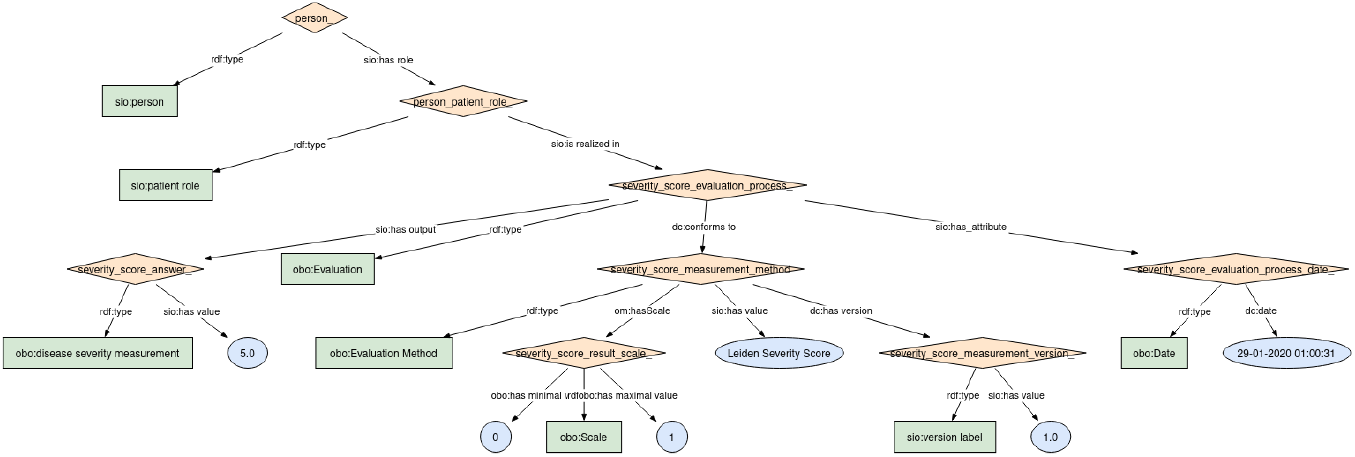
Semantic module to represent disease severity score phenotypes calculated in the hospital.

#### Ontological metadata model for COVID-19 resources

To allow the metadata of COVID-19 resources in the hospital to be findable, accessible, and reusable by both humans and machines, we provided an ontological model to expose it in a machine readable way. In practice, we designed a model by extending the DCAT2 based metadata model ^[1]^ that is to manage the metadata of common datasets. With four additional metadata elements from three standard ontologies, including the property “type” from the DCAT2, the properties “describes” and “data input of” from the Allotrope Foundation Ontologies (AFO) ^[2]^, and the property “has quality” from the OBO Relations Ontology (RO) ^[3]^, the metadata model features finer semantic granularity. In Figure 4, we show how we can specify that the BEAT-COVID data resource in our project is a *knowledge base*, that describes *COVID-19*, that is supposed to contain data input of *clinical studies*, and that has *synthetic quality* by means of these four object property values or edges in the RDF graph. This makes the structured semantics of the metadata of COVID-19 resources richer and more precise. The metadata model is publicly available on GitHub - metadata model.

**Figure 4.**
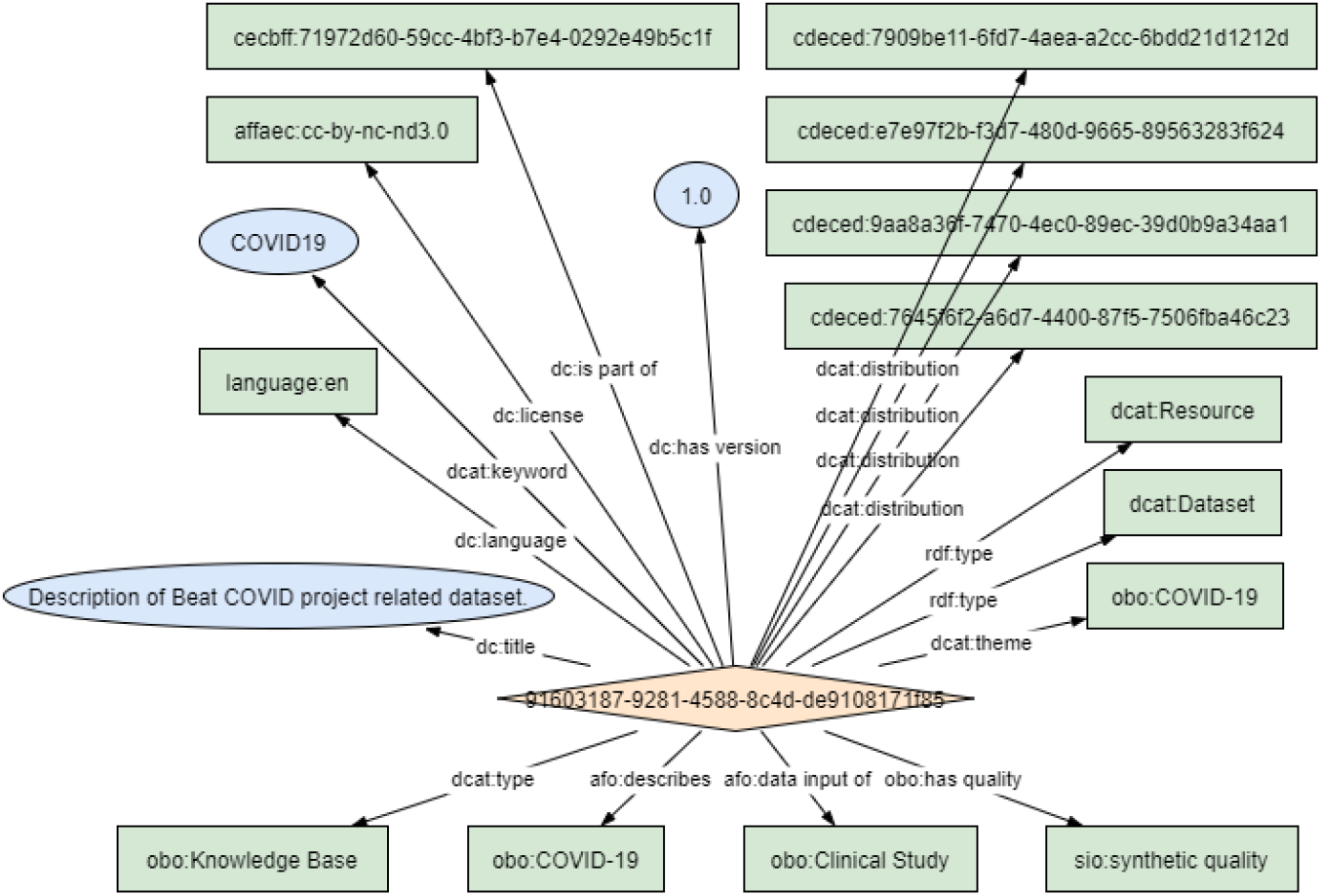
Ontological metadata model instantiated as an RDF graph. The four lower edges are the four additional metadata elements for COVID-19 data resource description.

#### FAIR Data Point for assessing the metadata of BEAT-COVID patient data

The basic idea of an FDP is to support scalable and transparent “routing” of data resources through stored metadata. The metadata stored and managed by an FDP makes the data resources described by the metadata semantically findable and reusable by machines. As an open gateway, it also makes different data resources accessible under defined constraints. Based on the designed ontological metadata model, we implemented an FDP to describe datasets in Opal and to publish FAIR metadata of these datasets on the Internet as complementary to the Mica system. This FDP publishes structured metadata for machines to automatically find BEATCOVID datasets and to interpret how to access and use the data stored in Opal, for instance to those algorithms visiting the data with the right access (Figure 5). Important to the FDP approach is that the data never leave its repository thereby protecting patient data and ensuring only authorized users have access. We performed an automated FAIR assessment of the same dataset from Mica described in the FDP. The results can be found here and showed that various aspects of the metadata description were improved in comparison to the Mica analysis results. For instance, FDP evaluation resulted in better identifier description of the (meta)data. With the publication of the BEAT-COVID resource metadata into the FDP we expect to increase the discoverability of COVID-19 patient data in the LUMC and to enable federated analytics for extended populations. To point out that an FDP is accessible and readable by machines through a REST API, and by humans through a Graphical User Interface (GUI). Note that the BEAT-COVID resource metadata is not all human readable. This is because the GUI of the current version of FDP only renders to the last fragment of a URI (Uniform Resource Identifier). For instance, the URI “www.example.org/ExOn/description” renders to the label “description” and the URI “www.example.org/ExOn/EL00001” renders to the label “EL 00001”. We are working on a more appropriate solution to display the “label” property from RDF Schema ^[4]^, following the best practice to always provide this label for humans. The FDP is publicly available at http://purl.org/biosemantics-lumc/beat-covid/fdp.

**Figure 5.**
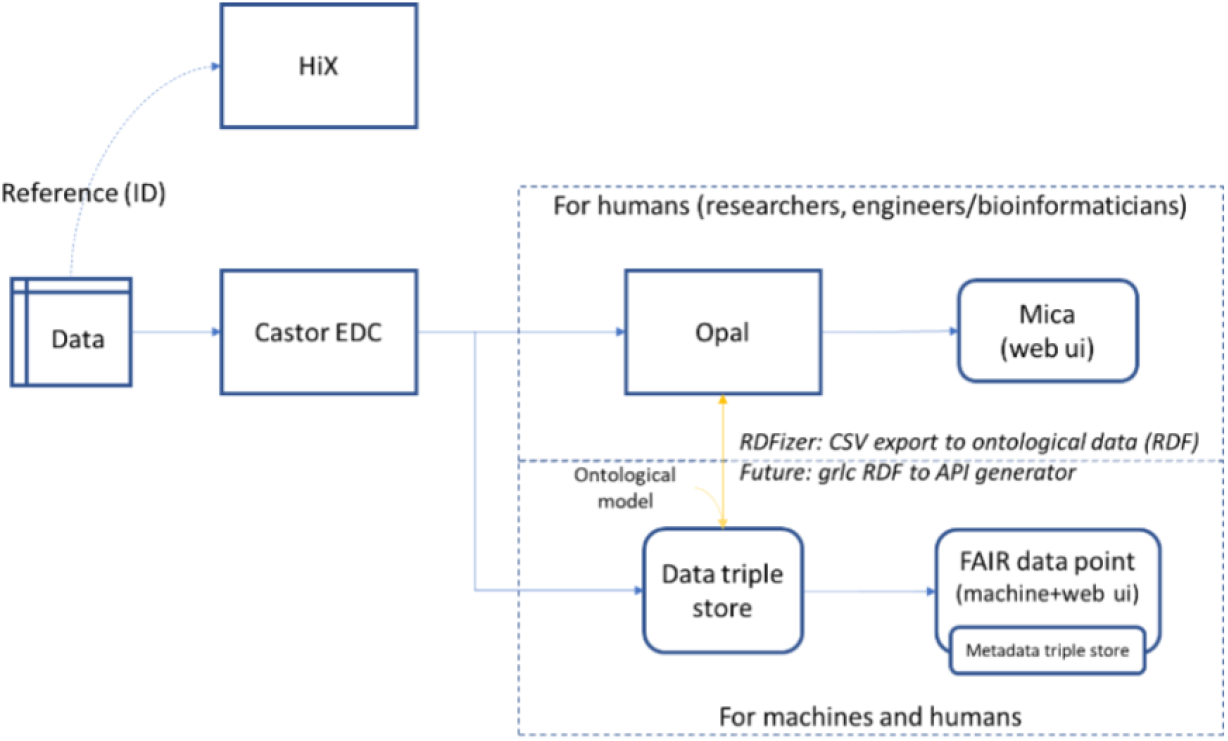
Integration of our ontological approach with existing systems.

#### Integrating the ontological models with the existing research data warehouse

Our next step was to add access to patient measurements as instances of the ontological model (‘ontologised data’) as a feature to the existing RDM. In Figure 5, we show how ontologised data is integrated with the existing Opal and Mica data management system. Our objective was to use the Opal and Mica systems as a foundation for FAIRification in the LUMC. While the Opal system manages integration of datasets in the hospital, the Mica system adds valuable metadata about the data resources. Even though Opal and Mica do not directly provide semantic modelling functionality, they do provide a basic annotation functionality that we used as the basis for connecting the ontological models. To instantiate the ontological linking data model in RDF, we developed an ‘RDFizer’ Python script as a minimal prototype for patient data FAIRification (see yellow arrow from Opal to Triple Store in Figure 5). Our current prototype uses CSV files with synthetic cytokine data as *input* to connect data from Opal to the ontological model that we developed for this data, thereby creating ‘ontologised data’ in RDF. Opal allows exporting datasets to CSV through its export function API ^[5]^.

Conversely, REST Web APIs can be generated from the ontologised data using the grlc server [37] (see yellow arrow from Triple Store to Opal in Figure 5). grlc is a tool to automatically convert SPARQL queries into REST Web APIs and make selected RDF data accessible to the Web. Moreover, it can translate SPARQL [38] queries stored and documented in GitHub repositories to Linked Data APIs on the fly. Essentially, it adds an additional communication layer that uses the common HTTP protocol on top of the SPARQL communication layer. To demonstrate this additional way of reusing FAIR patient data, we implemented a set of Web API endpoints to retrieve patient data in RDF. We first developed SPARQL queries, and then we ‘decorated’ and uploaded them in a GitHub repository - grlcqueries to be interpreted by grlc and build the API interface automatically. The SPARQL queries are examples of the potential power to execute sophisticated federated analysis that can be extended as more data resources become available. The Web API endpoints are publicly available at http://grlc.io/api-git/LUMC-BioSemantics/beat-covid-RESTful-API.

### Querying FAIR patient data with LOD for medical questions

To showcase that the FAIR RDM and the derived data infrastructure allow answering medical questions by querying patient data in terms of the ontological model and together with external open science knowledge, we performed two simple SPARQL queries on the synthetic cytokine data (Table 2). The queries were defined to answer the initial real world medical doctors’ hypothesis related to cytokines FAIR data. The first query demonstrates that clinical information from the LUMC can be queried, while the second demonstrates that queries can run across LUMC clinical data and external biomedical databases such as the UniProt protein knowledge-base by means of the federated SPARQL query shown in Figure 6. The SPARQL queries are available on GitHub - queries. The aforementioned grlc server provides an additional REST Web API for these queries.

**Table 2.** Example queries using external LOD resources.

| Question | Result |
| --- | --- |
| Count number of patients | LUMC query |
| Retrieve measured cytokines in the LUMC with protein annotation from the UniProt knowledgebase | Federated query |

**Figure 6.**
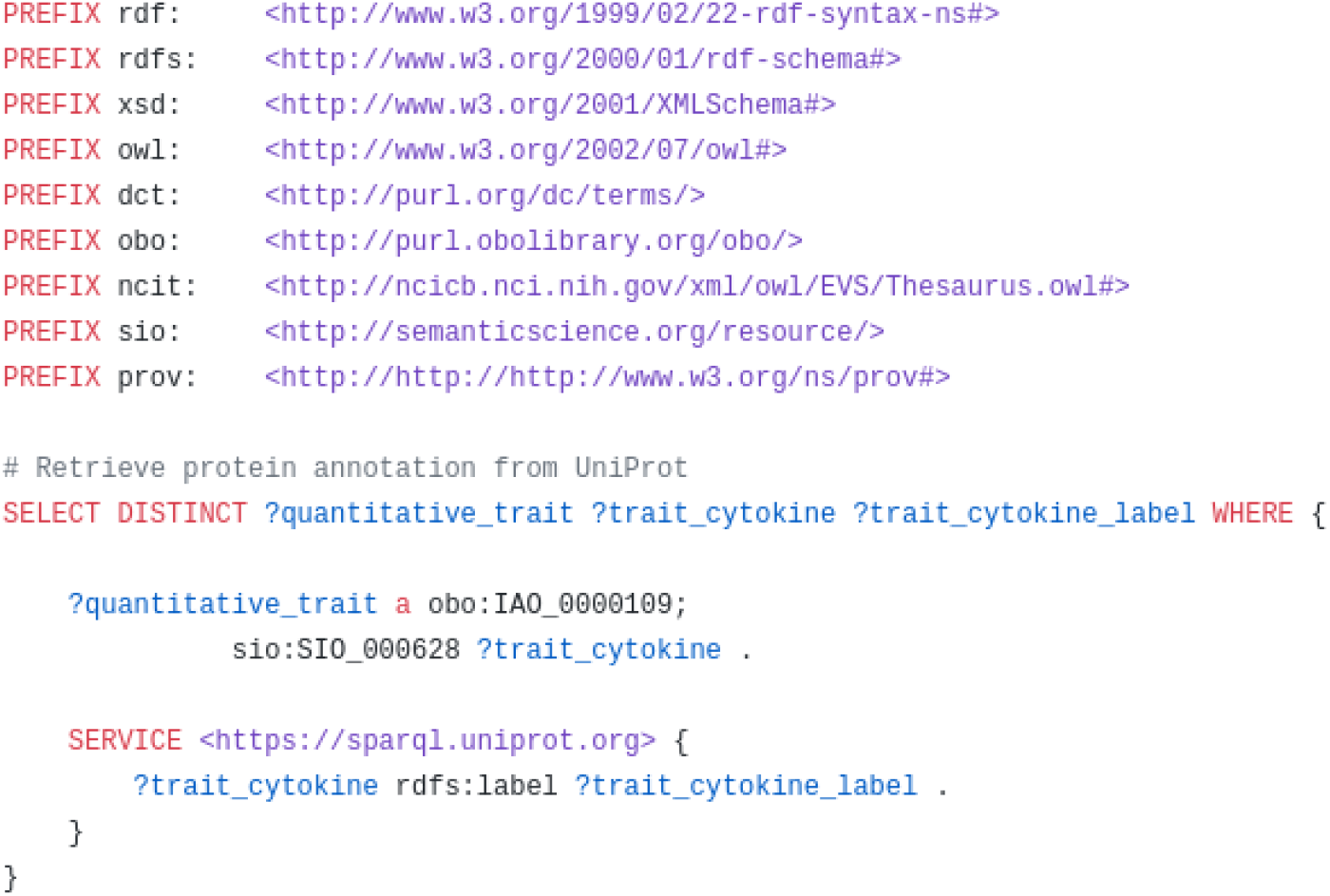
Federated SPARQL query crossing FAIR patient data with the UniProt knowledgebase.

## Discussion

### FAIRification in the hospital

The COVID-19 pandemic revealed how critically important it can be that patient data from multiple systems in the hospital are prepared for instant integrative analysis across those systems, as well as across hospitals and countries. This would be feasible if the hospital had a FAIR RDM plan that implied making patient data available as FDOs and thereby findable, accessible, interoperable, and reusable for computers [1]. However, COVID-19 patient data are not yet natively collected as FAIR data. Therefore, we have described a strategy to facilitate the adoption of the FAIR principles in the hospital based on the FAIR architecture shown in Figure 1 that complements an existing data management infrastructure. The strategy applies ontologies to increase the interoperability and machine readability of patient data records and patient datasets. We demonstrated that in the hospital (i) ontological models can complement existing data infrastructure, and (ii) they are an appropriate mechanism to formally capture agreement between stakeholders on what their data mean. They combine precise semantics for humans and corresponding actionable semantics for computers. Additional benefits are that they are extendible and they allow replacement with an improved ontological model (or adding multiple models). A similar ontology based approach is also applied to provide patient derived data as FDOs in biomedical and rare disease research such as in the EJP RD [21]. Interestingly, the results that we reused from the EJP RD project were addressing similar requirements as we had for COVID-19 data.

### Coordination with different stakeholders

The development of the FAIR RDM plan was made possible by a coordinated inter-disciplinary effort. In our experience, FAIRification requires at least data producers, data consumers, and FAIR data modellers [13, 39]. This is because the essential step of capturing the meaning of data in terms of ontologies requires the combined expertise of these stakeholders. In our case, this was available through the BEAT-COVID collaboration. The collaboration is providing user needs, technical requirements, insight in existing procedures and best practices regarding the management of the data lifecycle in the hospital. A clear challenge for our FAIRification process was communication between the different stakeholders with very different backgrounds.

This was further hampered by the communication limitations due to the pandemic itself. To mitigate the communication gap, we recorded meetings and shared material that was presented during the meetings. We also plan to organize Bring Your Own Data workshops to make stakeholders who are not FAIR experts more aware of the advantages that FAIR brings [40, 41, 42]. Under pressure of the urgency of the pandemic, we worked without dedicated FAIR stewards for this project. However, in going forward, this role seems essential to manage the necessary communication between disciplines [43].

### Establishing goals for FAIRification

Questions of researchers in the hospital were used as the drivers to establish FAIRification goals and to plan a FAIR RDM. The FAIRification preparation consisted of several meetings with medical doctors and clinical researchers. The focus of the meetings with domain experts was two-fold: (i) to identify the FAIRification goals, and (ii) to extract a set of specific research questions that drive the (meta)data modelling step. Both aims are related, because being able to answer *at least* the driving research questions is one of the main goals of FAIRification. The list of research questions included ‘What are the clinical parameters that can predict the disease course of a patient?’, ‘What are the biological pathways underlying patient symptoms and disease phenotypes?’, ‘How could biological pathways be positively or adversely affected by a particular treatment?’. The results of these meetings were guiding how data in the hospital should be interrelated and in what context they should be interpreted. We used this to define domain semantics in the context of testing and generating hypothesis with the help of OWL ontologies. The extendibility of ontologies mitigates the risk of limiting applications, because of initial overfitting on driving questions. Wider reusability of the FAIR RDM is a primary objective. To ensure that we are correctly capturing the semantics of knowledge and data, we are also exploring a formal method to validate the (meta)data models by the use of Competency Questions (CQs) and goal modelling. This will again rely on working with domain experts in close interdisciplinary collaboration. These research questions also facilitate communication between people of different expertise.

### Technical and social challenges and opportunities

For developing our approach within the BEAT-COVID collaboration, we took into account (i) the emergency of the situation, (ii) that various data management systems are in place at the hospital, (iii) that different types of data need to be prepared for timely exchange and efficient research. Consequently, our challenge was two-fold (i) to adapt our generic FAIRification workflow [12] in a hospital setting, (ii) to require minimal technical knowledge transfer, taking the opportunity of the combined expertise in the hospital that BEAT-COVID brought together. Key to our method is the development of two ontological models, one to enable analysis across clinical data (e.g. symptoms), investigational parameters (e.g. cytokine measurements), and data outside of the hospital, and another to represent the metadata of the patient data resources to increase the findability, accessibility and reusability. A metadata store was deployed conform to the FDP specification to provide access to this metadata. The metadata also includes a reference to access the ontological data. We demonstrated that Linked Data and Semantic Web technologies such as OWL ontologies, Triple Stores and the SPARQL query language provide the means to query patient data across sources in terms of the ontologies (Table 2). Taken together, these provide the FDOs for COVID-19 patient data and the basis for instant integrative federated analysis in the hospital.

While our ontological models aim to reflect our shared understanding of the data, a lack of tools still makes it challenging to transform health data to common data models such as HL7 FHIR [44], and for publishing it to findable resources [45]. There is a need for FAIRifier tools that support stakeholders in a clinical setting in every step from FAIR RDM planning to FAIR data creation, publication, evaluation, and reuse. Integration of FAIR implementations in existing data management tools such as Castor EDC can lower the burden substantially [39]. Similarly, the vocabulary and annotation features of Opal and Mica provide handles for future integration of FAIRification. The reuse of an abstract ontological data model, such as the EJP RD core model, in combination with the implementation of FDPs may further reduce thresholds for implementation and FAIR data reuse. An additional practical and technical challenge thereby is to protect patient identifying information but at the same time to have clinical data available close to real time. Classically most studies would retrieve data in retrospect from patient records. However, in the combat against COVID-19, first analyses were done when patients where still hospital admitted. Advanced data encryption was used to retrieve daily updates from patient records without including retrievable patient identifiers in the research data infrastructure. Although the big commitment of the BEAT-COVID group is facilitating the progress, other challenges for FAIRification in the hospital were ‘social’, presumably because stakeholders are not familiar with the steps that are needed to make a resource reusable by computers across multiple locations. We propose that a FAIR data policy is put in place for health research data conform [46]. To pave the way, there are several ongoing efforts to meet the need for education, such as FAIR training for researchers, clinicians and different types of stakeholders in organizations such as ELIXIR TeSS [47] and the EJP RD project for rare diseases.

### Patient data accessibility hurdles

Protecting patient data and privacy is a major concern and it is part of FAIRification to make a clear reference to how data are protected. As researchers, we must establish data management mechanisms that ensure that patient privacy is preserved and its usage under control. There are several options to deal with data privacy and safety such as using anonymised datasets, using substitute synthetic representations of sensitive datasets, and having the legal and ethical framework in place for the processing of sensitive personal data in the sense of the GDPR. As first step, the hospital needs to develop and implement a data governance policy that clearly specifies how to extract and apply the data as approved by the patient in the informed consent. Delaying data governance may delay the FAIRification process because it needs to be clear what data will be available and in which form to plan the FAIRification, but also to specify data governance in the metadata of the resource when an algorithm visits the data to use. Then, underdeveloped meta-data in data accessibility and data privacy hampers interoperability outside of the hospital. Consequently, it hampers data visiting, which means it hampers federated query and learning over FDPs and, therefore, limits hospital research capacity for analysis. Also, very important for accessibility and data privacy is that the digital objects *per se* can accommodate the criteria and protocols necessary to comply with regulatory and governance frameworks. Ontologies can aid in opening and protecting patient data by exposing logical definitions of data use conditions. Indeed, there are ontologies under development to define access and reuse conditions for patient data [24, 25]. Finally, it is worth noting that privacy preserving methods are available if data of the same person in multiple systems are required for a federated analysis [48, 49].

### International adoption of the FAIR principles for health data of hospitalised patients

The method for FAIRification that we described is focused on patient derived health data, down to the data record level. Two main outcomes are that we produced FAIR data for hospitalized patients, and we demonstrated that this data is instantly reusable for various secondary uses: for building software applications (and analysis workflows) via REST Web APIs, for querying cross-domain patient data and open public knowledge to add richer context to answer healthcare questions. While there are several projects that develop FAIRification procedures, they predominantly focus on life sciences data [14, 15, 50]. FAIR data in health is gaining momentum, and we already can find dedicated projects such as FAIR4Health [51] to use FAIR data in health to improve research. Our method has the same basis as the procedure followed earlier for rare disease patient registries (e.g. VASCA [13]), but here we integrated it with the hospital infrastructure, and demonstrated how the adoption of FAIR principles can be facilitated in the hospital through interdisciplinary collaboration. Hence, our experience may be valuable to national and global consensus on implementing FAIR principles in hospitals by the clinical community. For instance, the Dutch national Health Research Infrastructure (Health-RI) has stated that data stewardship at the Dutch University Medical Centres should adhere to the FAIR Principles [52]. Similar nationwide initiatives to improve health data reuse can be seen in Switzerland (Swiss Personalized Health Network [18]) and Germany (NFDI4Health [19]). These initiatives rely on a federated infrastructure, enhanced data interoperability and data linkage in compliance with privacy regulations for research. Our example has shown that FAIRification within the hospital can contribute to this infrastructure.

### Limitations and future work

We observed a number of limitations of our approach to enabling instant analysis of COVID-19 data across multiple hospital systems. First, we observed that the interdisciplinary collaboration and the willingness to implement FAIR principles, because of the pandemic, are not sufficient to provide easy access to data for implementing the FAIR services. A partial solution, at least to speed up the deployment of the FAIR services, could be to have synthetic patient data available. This could, for instance, be instantiated by Synthea [53, 54] from data in HL7 FHIR format. Second, at this time we have not incorporated a way to formally express patient consent and data usage conditions in our FAIR metadata. Currently, there are several efforts in human data communities to identify which elements are required, and standards are under development to capture these in machine readable ontological form, such as by the Informed Consent Ontology [24] and the Data Usage Ontology [25]. These can be linked into our FDP metadata model in the future. Third, we have not specifically addressed tooling (including standards) to support hospital data stewards in FAIR data management. This could pertain to tools for capturing FAIRification goals, ontological data modelling, data conversion, and mapping. Also, tools that evaluate the ‘FAIRness’ of data can guide the FAIRification process. This partly depends on the standards used by the domain of the data community providers [55], but it is not always clear what these standards are, if any. Current ongoing work in the FAIRification ‘world’ is to identify these community specific FAIR requirements and implementation choices. For instance, we envision as future work the establishment of FAIR maturity indicators for clinical data. Finally, we aim to progress on the opportunities for advancing research with FAIR patient data, further developing a FAIR Web API service to complement Opal APIs and knowledge graph based learning techniques. We would like to highlight the following developments.

#### Evaluation of ontological data models

We are evaluating the ontological models using CQs that are based on realistic questions posed by data model users [56], which are proposed as means to verify the scope (e.g., what is relevant to solve the challenges) and the relationships between concepts (e.g., check for missing or redundant relationships). A preliminary set of CQs from meetings with domain experts is available on GitHub - CQs.

#### COVID-19 hypothesis generation tool

We are developing a COVID-19 Hypothesis Generation tool for the LUMC based on the structured reviews for data and knowledge driven framework [57], as a means to exploit the FAIRification work for aiding medical doctors and researchers to answer their research questions. This framework has previously been used to support rare disease researchers to explore hypotheses as paths in case specific knowledge graph for their observations in the lab. After creating a preliminary knowledge graph with the FAIR synthetic cytokine data, we aim to incorporate background knowledge. The preliminary knowledge graph is available for browsing at LUMC BEAT-COVID Knowledge Graph.

#### Federated analytics across hospitals

We also aim to show how this FAIR infrastructure allows to query FAIR data from the BEAT-COVID project in the LUMC across other hospitals’ FAIR data without data leaving their source, i.e. the ‘data visiting’ approach. In the VODAN project, the GO FAIR VODAN in a box FDP [58] was used to test the trains and tracks of the PHT concept [59] and demonstrated the first intercontinental FDP SPARQL VODAN Africa proof of concept [60] developed by VODAN Africa and Asia - GO FAIR [2] query AllegroGraph WebView [61]. Secure FDP technology testing must be developed to implement trusted access control policies and to enable visiting synthetic datasets and pseudo-anonymised healthcare data. We aim to build on the VODAN and TWOC experiences and prepare an FDP instance that publishes BEAT-COVID metadata to be automatically found and used in trusted automated analytics workflows across multiple hospitals.

## Conclusion

We demonstrated that a FAIR research data management plan approach based on ontological models, open Science, Semantic Web technologies, and FDPs is a powerful method for generating FAIR patient data *at source*. FAIRification is providing data infrastructure that improves findability, accessibility, interoperability and reusability of patient real world observations in the hospital. Most importantly, we shown that FAIR patient data is machine actionable as digital objects linkable to LOD for analysis and ready to be used to develop applications for hypothesis generation and knowledge discovery on top. Finally, this work (in progress) showed what FAIRification entails in a real world hospital situation with existing infrastructure, different stakeholders and departments and the GDPR, and we discussed obstacles, challenges, solutions and future directions. We aim to provide a state of the art research data infrastructure in the hospital to deliver a federated solution enabling data access across the country and international borders, and accelerating research and translation to healthcare.

## Methods

We defined and implemented a method to make COVID-19 observational patient data in the hospital FAIR. This method is described in a detailed FAIRification workflow illustrated in Figure 7 and is an adapted version of the workflow presented by Jacobsen *et al*. [12]. We explicitly add the result obtained in each step, where applicable. We also include in which steps the FAIR experts worked in collaboration with other members of the BEAT-COVID group.

**Figure 7.**
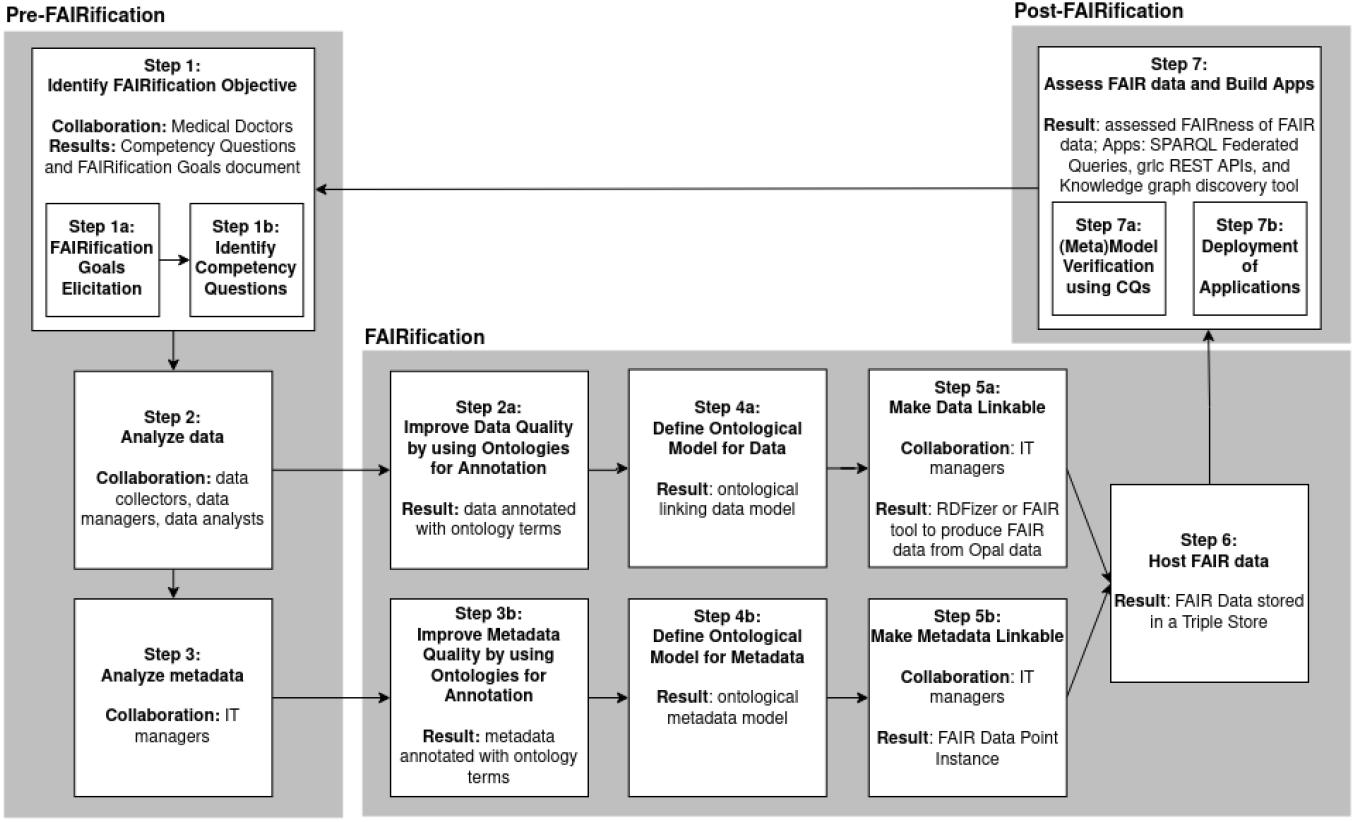
BEAT-COVID FAIRification workflow to make the data management and infrastructure in the hospital more FAIR. Collaborators and results are described in every step where applicable.

### Pre-FAIRification

#### Step 1: identify FAIRification objective

The first step was to determine the objective for making COVID-19 observational patient data FAIR in the hospital to define the specific FAIR requirements, implementations and workflow of this study. Medical doctors have pressing questions at point of care such as ‘What are the clinical parameters that can predict the disease course of a patient?’, ‘What are the biological pathways underlying patient symptoms and disease phenotypes?’, and ‘How can a patient be positively or adversely affected by a particular treatment?’. The FAIRification objective was therefore to prepare the diverse COVID-19 observational patient data to answer these questions. To this end, data needs to be integrated in a network and systems medicine approach [62], combined with external biomedical knowledge, and ready for computational analysis as illustrated in Figure 1.

#### Step 2 and step 3: analyze data and metadata

##### Research data management in the hospital

From admission date until discharge, patient data were collected by different departments. The types of COVID-19 observational data relevant for research, and so for FAIRification, were diverse: demographics information, clinical information, laboratory measurements, transcriptomics (RNA-Seq) data, metabolomics data, and if the patient was transferred to ICU, then data related to ICU outcome. The format depends on the different EDC systems used. Within LUMC, clinical and preclinical information were collected in HiX [63] and Castor EDCs [26], whereas ICU data was managed by the MetaVision software [64]. These EDC systems have different data access interfaces and use different technologies. To provide a single point of data access, research data were combined in the Opal data warehousing system. Opal is the OBiBa’s (Open Source Software for Epidemiology) core database application to store data in central data repositories that integrate under a uniform interface data collected from multiple sources, and it provides tools to import, transform and describe data [27]. Patient data was anonymised before importing it into Opal using advanced data encryption. Descriptions of the datasets, i.e metadata, stored in Opal were published on the Web through the Mica software application. Mica is used to create Web data portals for large scale studies or multiple study consortia. It provides a structured description of consortia, study catalogues and datasets, annotated and searchable data dictionaries, and data access request management. It is built upon a multitier architecture consisting of a REST application server for data management and administration, and clients to create and display data on the Web [28]. Opal and Mica are two standalone but interoperable software applications that provide features for management, harmonization, and analysis of epidemiological datasets [29, 65].

##### FAIR analysis of COVID-19 observational patient data

To improve the findability, accessibility, interoperability, and reusability of digital assets, we performed a FAIR analysis of (meta)data, i.e. an analysis of the FAIR status of data and metadata. We analysed data and databases to evaluate the FAIRification effort needed [12]. We started by analysing observational clinical measurements. We first got access to laboratory measurements of immunoresponse clinical parameters, cytokine levels, collected on different time points per patient to monitor its condition progress. Access to data was provided to us as an anonymised dataset. Then, we analysed the databases where these data were stored, which were first in Castor databases since this was the primary EDC system used in the hospital, second in Opal data warehouse since this system was used to integrate and store data from the various data sources. We investigated the representation (structure and format) and meaning (semantics) of the data, and the tools and technologies of each database system to optimize the FAIRification process of data.

### FAIRification

#### Step 2a and step 4a: improving interoperability with Semantic Web technologies and a linking data model

We described a synthetic cytokines dataset with ontologies. In Europe, GDPR imposes obligations onto organizations anywhere, so long as they target or collect data related to people in the EU. To comply with GDPR, we created a synthetic dataset of cytokine measurements, i.e. substituted synthetic representations of sensitive datasets, by using randomization for modelling patient data. This dataset contains basic information related to cytokine measurements and biosamples used per patient and time point, and a patient clinical identifier to link to clinical data. With the goals to answer research questions of medical doctors and make patient data machine readable to enable interoperability within data resources in the hospital and with external open science datasets such as LOD, we designed ontological models for cytokine lab measurements, biosamples and severity scores to represent data based on the Linked Data principles [66] and Semantic Web technologies such as the W3C recommended RDF and OWL standards [32, 31]. Our approach was to define a conceptual model as an abstract and reusable model to capture as much of patient data (measurements, biosamples and score phenotypes), by using standard common schemas and well established ontologies and vocabularies widely used by the biomedical community such as the ones in the OBO Foundry [30]. With this approach we created an ontological linking model for cytokines measurements dataset from the laboratory.

#### Step 3a and step 4b: improving findability, accessibility, interoperability and reusability with Semantic Web technologies, a metadata model and FAIR Data Points

With the goals to answer research questions of medical doctors and make resource metadata human and machine readable to enable cross-resource data analytics, we designed a metadata ontological model and implemented an FDP instance [23] to make LUMC COVID-19 digital objects findable for machines on the Internet. An FDP is a Web application that enables data owners to expose information about their datasets using rich machine actionable metadata. It allows creating, storing, and serving FAIR metadata about datasets and its distributions for both humans and machines. An FDP does *not* enable open access, but the metadata is expected to include information about what the resource contains and how datasets and content can be accessed under defined conditions. Opening up FAIR (meta)data by publishing them on an FDP allows algorithms to search these (meta)data, looking for patterns [67]. Mica is a tool to expose datasets from an Opal database on the Internet through Web portals that allow (meta)data descriptions. An FDP provides additional means to expose FAIR metadata, i.e. machine actionable, via the FDP specification, a standardized metadata ontological model based on DCAT [22]. FDP also exposes (meta)data via a REST Web API that enables client applications to automate retrieval, aggregation and filtering (meta)data from distributed FDPs. We used FDP v1.10.0.

#### Step 5 and step 6: make (meta)data as Linked Data and host FAIR data

To host and publish patient data, we cut the original synthetic cytokine patient dataset into a few rows. We generated patient Linked Data using this synthetic patient data we created as *input* and instantiating the linking data ontological model. To do it we developed ‘RDFizer’ a FAIRification tool in Python 3 that parses and converts the synthetic data CSV file into RDF. To host the generated FAIR data, we used the free edition of GraphDB Triple Store [68] v9.7.0 where the data is natively stored as RDF. We implemented an FDP instance where the metadata ontological model is described and published as DCAT based Linked Data.

### Post-FAIRification

#### Step 7: assessment and software applications

##### Evaluation

We evaluated the discoverability of the BEAT-COVID resource by means of the FAIR Maturity Indicators evaluator tool [69]. We have evaluated our ontological models by means of several CQs [56] (in progress). We have answered the questions using SPARQL queries for the sake of reusability, then users can reuse the queries if they want updated answers in the future.

##### Data analytics with Semantic Web technologies

We used the W3C recommended SPARQL query language [38] to perform data analytics over the LUMC RDF patient data and across diverse external data sources in LOD. We used free edition of GraphDB Triple Store v9.7.0 for our use case where the data is natively stored as RDF.

##### Web API development

We used grlc version 1.3.6 [37] to enable programmatic access to FAIR data in the hospital. Grlc is a lightweight server that automatically builds consistent, well documented and neatly organized Linked Data APIs on the fly, with no input required from users beyond a URL path to a GitHub repository hosting a set of SPARQL queries that complies with the specific grlc syntax ^[6]^. It provides three basic operations: 1. generates the Swagger spec of a specified GitHub repository; 2. generates the Swagger UI to provide an interactive user facing frontend of the API contents; and 3. translates SPARQL queries into HTTP requests to call the operations of the API against a SPARQL endpoint with parameters set in the queries.

##### Hypothesis generation tool

We used the Neo4j graph database framework [70] as used in the structured reviews approach [57] for storage, management and mining of FAIR patient data. The graph database technology has been shown to facilitate management and exploration of biomedical knowledge [71]. Neo4j graph database enables users to query the knowledge graph using the Cypher query language, either through an API or a GUI. RDF data was imported into the Neo4j Community Server v4.2.5 graph database through the Neo4j neosemantics toolkit v4.2.0 [72].

## Data Availability

All data and code are publicly available under license terms.

https://github.com/LUMC-BioSemantics/beat-covid

## Appendix

## Acknowledgements

We would like to specially thank Eleni Mina, Tooba Abassi-Daloii, Daniël Wijnbergen, Winette Koning, Luiz Olavo Bonino da Silva and Katy Wolstencroft. We would also like to thank our EJP RD colleagues Peter-Bram ‘t Hoen and Mark Wilkinson for all the discussions. Finally, we would like to thank Professor Barend Mons for inspiring us to make a real difference in data sharing and knowledge representation.

## Abbreviations

FAIR: Findable, Accessible, Interoperable and Reusable
VODAN: Virus Outbreak Data Network
TWOC: Trusted World of Corona
FDO: FAIR Digital Object
GDPR: General Data Protection Regulation
LUMC: Leiden University Medical Center
RDM: Research Data Management
EJP RD: European Joint Programme Rare Diseases
DCAT: Data Catalogue Vocabulary
FDP: FAIR Data Point
LOD: Linked Open Data
EDC: Electronic Data Capture
OBO: Open Biological Biomedical Ontologies
OWL: Web Ontology Language
RDF: Resource Description Framework
ICU: Intensive Care Unit
GUI: Graphical User Interface
URI: Uniform Resource Identifier
CQ: Competency Question.

## Availability of data and materials

The datasets supporting the conclusions of this article are available in the following repositories. The ontological models, the SPARQL queries, grlc SPARQL queries, SPARQL CQs and scripts are freely available at the Biosemantics (GitHub): The data model is available at https://github.com/LUMC-BioSemantics/beat-covid/tree/master/fair-data-model/cytokine/model-triples

The metadata model is available at https://github.com/LUMC-BioSemantics/beat-covid/tree/master/fair-metadata-model

Synthetic cytokine patient dataset in CSV is available at https://github.com/LUMC-BioSemantics/beat-covid/tree/master/fair-data-model/cytokine/synthetic-data

Source code for RDFizer is available at https://github.com/LUMC-BioSemantics/beat-covid/tree/master/fair-data-model/scripts/rdfizer

COVID-19 synthetic patient cytokine knowledge graph in RDF is available at https://github.com/LUMC-BioSemantics/beat-covid/tree/master/fair-data-model/cytokine/rdf

RDF data is accessible through the LUMC Beat-COVID FDP at https://w3id.org/biosemantics-lumc/beat-covid/fdp/

Source code for FDP implementation is freely available at the FAIRDataPoint at https://github.com/FAIRDataTeam/FAIRDataPoint

SPARQL queries are available at https://github.com/LUMC-BioSemantics/beat-covid/tree/master/fair-data-model/cytokine/sparql-queries grlc endpoint APIs are available at http://grlc.io/api-git/LUMC-BioSemantics/beat-covid-RESTful-API grlc SPARQL queries are available at https://github.com/LUMC-BioSemantics/beat-covid-RESTful-API

Evaluations: FAIR assessment results of a dataset described in Mica are available at https://fairsharing.github.io/FAIR-Evaluator-FrontEnd/#!/evaluations/4081, and the FAIR assessment results of the same dataset, but described in a FDP are available at https://fairsharing.github.io/FAIR-Evaluator-FrontEnd/#!/evaluations/5589

SPARQL CQs are available at https://github.com/LUMC-BioSemantics/beat-covid/tree/master/fair-data-model/cytokine/competency-questions

Figures: All model figures both in this manuscript and in GitHub project repository were automatically produced using the corresponding RDF/Turtle file as *input* and the Web drawing tool at https://w3id.org/ejp-rd/tools/rdf-drawing

## Ethics approval and consent to participate

Not applicable.

## Consent for publication

Not applicable.

## Authors’ contributions

MR conceived the initial RDM plan. The BEAT-COVID group provided feedback and guidance on research priorities, the meaning of data types, and the RDM plan. NQR, RK, CHB, QL and AJ conceptualised and realised the RDM plan. NQR, CHB, QL, AJ and RK contributed in regular FAIRification discussions. SJ provided guidance and access to the laboratory measurements data. The COVID-19 LUMC group provided data to the BEAT-COVID project. HJW provided guidance and access to the Opal and Mica software applications used in the existing research data management. RK, QL, KB and ELAF contributed to the harmonization of the BEAT-COVID FDP with the LUMC VODAN FDP; NQR drafted the initial version of the manuscript. AJ and MR revised the manuscript. MR, BM and the BEAT-COVID group acquired funding to support this work. All authors reviewed and approved the final version of the manuscript.

## Authors’ information

### BEAT-COVID group (in alphabetical order, IR)

M. Sesmu Arbous^1^, Bernard M. van den Berg^2^, Suzanne Cannegieter^3^, Christa M. Cobbaert^4^, Anne M. van der Does^5^, Jacques J.M. van Dongen^6^, Jeroen Eikenboom^7^, Mariet C.W. Feltkamp^8^, Annemieke Geluk^9^, Jelle J. Goeman^10^, Martin Giera^11^, Thomas Hankemeier^12^, Mirjam H.M. Heemskerk^13^, Pieter S. Hiemstra^5^, Cornelis H. Hokke^14^, Jacqueline J. Janse^14^, Simon P. Jochems^14^, Simone A. Joosten^9^, Marjolein Kikkert^8^, Lieke Lamont^12^, Judith Manniën^10^, Tom H.M. Ottenhoff^9^, T. Pongracz^11^, Michael R. del Prado^1^, Núria Queralt-Rosinach^15^, Meta Roestenberg^9,14^, M. Roos^15^, Anna H.E. Roukens^9^, Hermelijn H. Smits^14^, Eric J. Snijder^8^, Frank J.T. Staal^6^, Leendert A. Trouw^6^, Roula Tsonaka^10^, Aswin Verhoeven^11^, Leo G. Visser^9^, Jutte J.C. de Vries^8^, David J. van Westerloo^1^, Jeanette Wigbers^1^, Henk J. van der Wijk^10^, Robin C. van Wissen^4^, Manfred Wuhrer^11^, Maria Yazdanbakhsh^14^, Mihaela Zlei^6^

1. Dept. of Intensive Care, LUMC
2. Dept. of Internal Medicine, LUMC
3. Dept. of Clinical Epidemiology, LUMC
4. Dept. of Clinical Chemistry, LUMC
5. Dept. of Pulmonology, LUMC
6. Dept. of Immunology, LUMC
7. Dept. of Internal Medicine, LUMC
8. Dept. of Medical Microbiology, LUMC
9. Dept. of Infectious Diseases, LUMC
10. Dept. of Biomedical Data Sciences, LUMC
11. Center for Proteomics and Metabolomics, LUMC
12. Division of Systems Biomedicine and Pharmacology, Leiden Academic Center for Drug Research, Leiden University, the Netherlands
13. Dept. of Hematology, LUMC
14. Dept. of Parasitology, LUMC
15. Dept. of Human Genetics, LUMC

### COVID-19 LUMC group (IR)

Josine A. Oud, MSc^1^; Meryem Baysan, MSc^2^, 3; Jeanette Wigbers^2^; Lieke J. van Heurn, BSc^3^; Susan B. ter Haar, BSc^3^; Alexandra G.L. Toppenberg, BSc^3^; Laura Heerdink, BSc^3^; Annekee A. van IJlzinga Veenstra, BSc^3^; Anna M. Eikenboom, BSc^3^; Julia M. Wubbolts, MSc^4^; Jonathan Uzorka MD^4^, Willem Lijfering MD PhD^3^; Romy Meier^1^; Ingeborg de Jonge^3^; Sesmu M. Arbous MD PhD^2^; Mark G.J. de Boer MD PhD^4^; Anske G. van der Bom, MD PhD^3^; Olaf M. Dekkers, MD PhD^3^: Frits Rosendaal, MD PhD^3^

1. Dept. of Hematology, LUMC
2. Dept. of Intensive Care, LUMC
3. Dept. of Clinical Epidemiology, LUMC
4. Dept. of Infectious Diseases, LUMC

## Footnotes

[1] https://www.w3.org/TR/vocab-dcat-2/

[2] https://www.allotrope.org/ontologies

[3] http://www.obofoundry.org/ontology/ro.html

[4] https://www.w3.org/TR/rdf-schema/

[5] https://opaldoc.obiba.org/en/latest/python-user-guide/export/export-csv.html

[6] https://github.com/CLARIAH/grlc

## References

1. Wilkinson, M.D., Dumontier, M., Aalbersberg, I.J., Appleton, G., Axton, M., Baak, A., Blomberg, N., Boiten, J.-W., da Silva Santos, L.B., Bourne, P.E., et al.: The FAIR guiding principles for scientific data management and stewardship. Scientific data 3 (2016)

2. GO FAIR: Virus Outbreak Data Network (VODAN) (2021). https://www.go-fair.org/implementation-networks/overview/vodan/ Accessed Accessed 23 Jul 2021

3. ZonMw: COVID-19 Programme (2021). https://www.zonmw.nl/en/research-and-results/infectious-diseases-and-antimicrobial-resistance/programmas/programme-detail/covid-19-programme/ Accessed Accessed 23 Jul 2021

4. Health Holland: Trusted World of Corona (TWOC) (2021). https://www.health-holland.com/project/2020/trusted-world-of-corona Accessed Accessed 23 Jul 2021

5. ELIXIR: ELIXIR COVID-19 Services (2021). https://elixir-europe.org/services/covid-19 Accessed Accessed 27 Jul 2021

6. Luiz Olavo Bonino da Silva Santos: FAIR Digital Object Framework (2020). https://fairdigitalobjectframework.org/ Accessed Accessed 27 Jul 2021

7. Lamprecht, A.L., Garcia, L., et al.: Towards fair principles for research software. Data Science 3, 37–59 (2020)

8. GO FAIR: Data Together COVID-19 Appeal and Actions (2020). https://www.go-fair.org/wp-content/uploads/2020/03/Data-Together-COVID-19-Statement-FINAL.pdf Accessed Accessed 23 Jul 2021

9. van Soest J, C,S., O, M., et al.: Using the personal health train for automated and privacy-preserving analytics on vertically partitioned data. Studies in health technology and informatics 247, 581–585 (2018)

10. van Soest; Oliver Kohlbacher; Lukas Zimmermann; Holger Stenzhorn; Md. Rezaul Karim; Michel Dumontier; Stefan Decker; Luiz Olavo Bonino da Silva Santos; Andre Dekker, O.B.A.C.J.: Distributed analytics on sensitive medical data: The personal health train. Data Intelligence 2, 96–107 (2020). doi:10.1162/dinta00032

11. Landi, A., Thompson, M., Giannuzzi, V., Bonifazi, F., Labastida, I., da Silva Santos, L.O.B., Roos, M.: The “A” of FAIR – As Open as Possible, as Closed as Necessary. Data Intelligence 2(1-2), 47–55 (2020). doi:10.1162/dinta00027. https://direct.mit.edu/dint/article-pdf/2/1-2/47/1893372/dinta00027.pdf

12. da Silva Santos; Barend Mons; Erik Schultes; Marco Roos; Mark Thompson, A.J.R.K.L.O.B.: A generic workflow for the data fairification process. Data Intelligence 2, 56–65 (2020). doi:10.1162/dinta00028

13. Groenen, K.H.J., Jacobsen, A., Kersloot, M.G., Vieira, B., van Enckevort, E., Kaliyaperumal, R., Arts, D.L., ‘t Hoen, P.A.C., Cornet, R., Roos, M., Kool, L.S.: The de novo fairification process of a registry for vascular anomalies. medRxiv (2020). doi:10.1101/2020.12.12.20245951. https://www.medrxiv.org/content/early/2020/12/14/2020.12.12.20245951.full.pdf

14. FAIRplus project: FAIR Cookbook. The FAIR Cookbook: a deliverable of the FAIRplus project (grant agreement 802750), funded by the IMI programme, a private-public partnership that receives support from the European Union’s Horizon 2020 research and innovation programme and EFPIA Companies. (2019). https://fairplus.github.io/the-fair-cookbook/content/home.html Accessed Accessed 26 Jul 2021

15. Innovative Medicine Initiative: FAIRplus project (2019). https://fairplus-project.eu/ Accessed Accessed 26 Jul 2021

16. GO FAIR VODAN: A three-point framework for FAIRification (2020). https://www.go-fair.org/2020/07/08/a-three-point-framework-for-fairification/ Accessed Accessed 28 Jul 2021

17. dos Santos Vieira, B., et al.: A de novo fairification process for rare disease registries. In: Abstracts of the International Congress of Research on Rare and Orphan Diseases: January 13-15, 2021; Online, p. 67 (2021). https://www.react-congress.org/wp-content/uploads/Abstract_book_REACTxIRDIRCongress_2021.pdf

18. Swiss Academy of Medical Sciences: Swiss Personalized Health Network (2020). https://sphn.ch/ Accessed Accessed 28 Jul 2021

19. NFDI4Health: NFDI4Health Nationale Forschungsdateninfrastruktur für personenbezogene Gesundheitsdaten (2021). https://www.nfdi4health.de/ Accessed Accessed 28 Jul 2021

20. Roukens, A.H.E., König, M., Dalebout, T., Tak, T., Azimi, S., Kruize, Y., Pothast, C.R., Hagedoorn, R.S., Arbous, S.M., Zhang, J.L.H., Verheij, M., Prins, C., van der Does, A.M., Hiemstra, P.S., de Vries, J.J.C., Janse, J.J., Roestenberg, M., Myeni, S.K., Kikkert, M., Heemskerk, M.H.M., Yazdanbakhsh, M., Smits, H.H., Jochems, S.P., group, B.-C.: Prolonged activation of nasal immune cell populations and development of tissue-resident sars-cov-2 specific cd8 t cell responses following covid-19. medRxiv (2021). doi:10.1101/2021.04.19.21255727. https://www.medrxiv.org/content/early/2021/04/22/2021.04.19.21255727.full.pdf

21. Kaliyaperumal, R., Wilkinson, M.D., Alarcón Moreno, P., Benis, N., Cornet, R., dos Santos Vieira, B., Dumontier, M., Bernabé, C.H., Jacobsen, A., Le Cornec, C.M.A., Godoy, M.P., Queralt-Rosinach, N., Schultze Kool, L.J., Swertz, M.A., van Damme, P., van der Velde, K.J., van Lin, N., Zhang, S., Roos, M.: Semantic modelling of common data elements for rare disease registries, and a prototype workflow for their deployment over registry data. medRxiv (2021). doi:10.1101/2021.07.27.21261169. https://www.medrxiv.org/content/early/2021/07/30/2021.07.27.21261169.full.pdf

22. W3C: DCAT2 W3C Homepage (2020). https://www.w3.org/TR/vocab-dcat-2/ Accessed Accessed 24 Aug 2020

23. da Silva Santos, L.O.B., Wilkinson, M.D., Kuzniar, A., Kaliyaperumal, R., Thompson, M., Dumontier, M., Burger, K.: Fair data points supporting big data interoperability. London: ISTE Press 4, 270–279 (2016)

24. Lin, Y., Harris, M., Manion, F., Eisenhauer, E., Zhao, B., Shi, W., Karnovsky, A., He, Y.: Development of a bfo-based informed consent ontology (ico). In: ICBO (2014)

25. EMBL-EBI: The Data Use Ontology (DUO) (2021). https://github.com/EBISPOT/DUO Accessed Accessed 28 Jul 2021

26. castor: Castor Homepage (2021). https://www.castoredc.com/ Accessed Accessed 20 Aug 2020

27. OBiBa: Opal OBiBa’s software Homepage (2020). https://www.obiba.org/pages/products/opal/ Accessed Accessed 20 Aug 2020

28. OBiBa: Mica OBiBa’s software Homepage (2020). https://www.obiba.org/pages/products/mica/ Accessed Accessed 20 Aug 2020

29. Doiron DF.I.B.P.F.V. Marcon Y: Software application profile: Opal and mica: open-source software solutions for epidemiological data management, harmonization and dissemination. Int J Epidemiol. 46(5), 1372–1378 (2017)

30. Smith, B., Ashburner, M., Rosse, C., Bard, J., Bug, W., Ceusters, W., Goldberg, L.J., Eilbeck, K., Ireland, A., Mungall, C.J., Leontis, N., Rocca-Serra, P., Ruttenberg, A., Sansone, S.-A., Scheuermann, R.H., Shah, N., Whetzel, P.L., Lewis, S.: The OBO Foundry: Coordinated Evolution of Ontologies to Support Biomedical Data Integration. Nature Biotechnology 25(11), 1251–1255 (2007). doi:10.1038/nbt1346

31. OWL W3C Homepage. https://www.w3.org/2001/sw/wiki/OWL Accessed Accessed 21 Aug 2020

32. RDF W3C Homepage. https://www.w3.org/RDF/ Accessed Accessed 21 Aug 2020

33. EJP RD: EJP RD core CDE semantic model. https://github.com/ejp-rd-vp/CDE-semantic-model/wiki/Core-model-SIO Accessed Accessed 18 Oct 2020

34. Queralt-Rosinach, N., Bello, S., Hoehndorf, R., Weiland, C., Rocca-Serra, P., Schofield, P.N.: Modeling quantitative traits for covid-19 case reports. medRxiv (2020). doi:10.1101/2020.06.18.20135103. https://www.medrxiv.org/content/early/2020/06/20/2020.06.18.20135103.full.pdf

35. Acute Physiology And Chronic Health Evaluation (APACHE): APACHE IV Score (2021). https://intensivecarenetwork.com/Calculators/Files/Apache4.html Accessed Accessed 28 Jul 2021

36. Sequential Organ Failure Assessment (SOFA): SOFA Score (2021). https://www.mdcalc.com/sequential-organ-failure-assessment-sofa-score Accessed Accessed 28 Jul 2021

37. Meronó-Peñuela A. H.R.: grlc makes github taste like linked data apis. In: Sack H., S.N.M.D.A.S.L.C. Rizzo G. (ed.) The Semantic Web. ESWC 2016. Lecture Notes in Computer Science, Vol 9989 (2016). https://doi.org/10.1007/978-3-319-47602-548

38. SPARQL W3C Homepage. https://www.w3.org/TR/rdf-sparql-query/ Accessed Accessed 21 Aug 2020

39. Kersloot, M.G., Jacobsen, A., Groenen, K.H.J., Vieira, B.d.S., Kaliyaperumal, R., Abu-Hanna, A., Cornet, R., ‘t Hoen, P.A.C., Roos, M., Kool, L.S., Arts, D.L.: De-novo fairification via an electronic data capture system by automated transformation of filled electronic case report forms into machine-readable data. medRxiv (2021). doi:10.1101/2021.03.04.21250752. https://www.medrxiv.org/content/early/2021/03/08/2021.03.04.21250752.full.pdf

40. M, R., P, L.: Bring your own data parties and beyond: make your data linkable to speed up rare disease research. Rare Diseases and orphan drugs An International Journal of Public Health 1(4), 21 (2014)

41. M, R., et al.: Bring your own data workshops: a mechanism to aid data owners to comply with linked data best practices. In: Paschke, A., Burger, A., Romano, P., Marshall, M.S., Splendiani, A. (eds.) Proceedings of the 7th International Workshop on Semantic Web Applications and Tools for Life Sciences: December 9-11, 201; Berlin, Germany, pp. 16–27 (2014). http://ceur-ws.org/Vol-1320/paper_36.pdf

42. ELIXIR-EXCELERATE: Bring Your Own Data (2019). https://doi.org/10.5281/zenodo.3207809 Accessed Accessed 28 Jul 2021

43. Salome Scholtens, Mijke Jetten, Jasmin Böhmer, Christine Staiger, Inge Slouwerhof, Marije van der Geest, & Celia W.G. van Gelder: Final report: Towards FAIR data steward as profession for the lifesciences. Report of a ZonMw funded collaborative approach built on existing expertise (2019). https://doi.org/10.5281/zenodo.3474630 Accessed Accessed 5 Aug 2021

44. HL7 International: HL7 FHIR Homepage (2019). https://www.hl7.org/fhir/ Accessed Accessed 18 Oct 2020

45. Löbe, M., Matthies, F., Stäubert, S., Meineke, F.A., Winter, A.: Problems in fairifying medical datasets. In: Pape-Haugaard, L.B., Lovis, C., Madsen, I.C., Weber, P., Nielsen, P.H., Scott, P. (eds.) Digital Personalized Health and Medicine - Proceedings of MIE 2020, Medical Informatics Europe, Geneva, Switzerland, April 28 - May 1, 2020. Studies in Health Technology and Informatics, vol. 270, pp. 392–396 (2020). doi:10.3233/SHTI200189. https://doi.org/10.3233/SHTI200189

46. FAIR4Health: D2.3. Guidelines for implementing FAIR Open Data policy in health research (2019). https://www.fair4health.eu/en/resources/project-deliverable Accessed Accessed 24 Ago 2020

47. ELIXIR: ELIXIR’s Training Portal (2020). https://tess.elixir-europe.org/ Accessed Accessed 28 Jul 2021

48. A. Hörbst, G.S. D. Hayn: EHealth2014 – Health Informatics Meets EHealth, (2014)

49. Baker, D.B., Knoppers, B.M., Phillips, M., van Enckevort, D., Kaufmann, P., Lochmuller, H., Taruscio, D.: Privacy-preserving linkage of genomic and clinical data sets. IEEE/ACM Transactions on Computational Biology and Bioinformatics 16(4), 1342–1348 (2019). doi:10.1109/TCBB.2018.2855125

50. ELIXIR: RDMkit (2021). https://rdmkit.elixir-europe.org/ Accessed Accessed 26 Jul 2021

51. FAIR4Health: FAIR4Health (2019). https://www.fair4health.eu/ Accessed Accessed 28 Jul 2021

52. Health-RI: FAIR Principles (2019). https://www.health-ri.nl/initiatives/personal-health-train Accessed Accessed 28 Jul 2021

53. Walonoski, J., Kramer, M., Nichols, J., Quina, A., Moesel, C., Hall, D., Duffett, C., Dube, K., Gallagher, T., McLachlan, S.: Synthea: An approach, method, and software mechanism for generating synthetic patients and the synthetic electronic health care record. Journal of the American Medical Informatics Association 25(3), 230–238 (2017). doi:10.1093/jamia/ocx079. https://academic.oup.com/jamia/article-pdf/25/3/230/34150150/ocx079.pdf

54. MITRE: Synthea (2021). https://synthetichealth.github.io/synthea/ Accessed Accessed 28 Jul 2021

55. Jacobsen, A., de Miranda Azevedo, R., Juty, N., Batista, D., Coles, S., Cornet, R., Courtot, M., Crosas, M., Dumontier, M., Evelo, C.T., Goble, C., Guizzardi, G., Hansen, K.K., Hasnain, A., Hettne, K., Heringa, J., Hooft, R.W.W., Imming, M., Jeffery, K.G., Kaliyaperumal, R., Kersloot, M.G., Kirkpatrick, C.R., Kuhn, T., Labastida, I., Magagna, B., McQuilton, P., Meyers, N., Montesanti, A., van Reisen, M., Rocca-Serra, P., Pergl, R., Sansone, S.-A., da Silva Santos, L.O.B., Schneider, J., Strawn, G., Thompson, M., Waagmeester, A., Weigel, T., Wilkinson, M.D., Willighagen, E.L., Wittenburg, P., Roos, M., Mons, B., Schultes, E.: FAIR Principles: Interpretations and Implementation Considerations. Data Intelligence 2(1-2), 10–29 (2020). doi:10.1162/dint_r_00024. https://direct.mit.edu/dint/article-pdf/2/1-2/10/1893430/dint_r_00024.pdf

56. Grüninger, M., Fox, M.S.: Methodology for the design and evaluation of ontologies. (1995)

57. Queralt-Rosinach, N., Stupp, G.S., Li, T.S., Mayers, M., Hoatlin, M.E., Might, M., Good, B.M., Su, A.I.: Structured reviews for data and knowledge-driven research. Database 2020 (2020). doi:10.1093/database/baaa015.baaa015. https://academic.oup.com/database/article-pdf/doi/10.1093/database/baaa015/33034124/baaa015.pdf

58. GO FAIR: VODAN in a Box: the all in one solution for easy instalment of VODAN FAIR Data Points (2020). https://www.gofairfoundation.org/vodan-in-a-box-the-all-in-one-solution-for-easy-instalment-of-vodan-fair-data-points/ Accessed Accessed 28 Jul 2021

59. Health-RI: Personal Health Train (2019). https://www.health-ri.nl/initiatives/personal-health-train Accessed Accessed 28 Jul 2021

60. GO FAIR: Proof of Concept developed by VODAN Africa and Asia (2020). https://www.go-fair.org/2020/10/27/proof-of-concept-developed-by-vodan-africa-and-asia/ Accessed Accessed 28 Jul 2021

61. GO FAIR: AllegroGraph WebView (lumc.nl) (2020). https://sparql.lumc.nl:8443/#/repositories/crf/query/r/demo-query Accessed Accessed 28 Jul 2021

62. Comte B, B.A.e.a. Baumbach J: Network and systems medicine: Position paper of the european collaboration on science and technology action on open multiscale systems medicine. Network and systems medicine 3(1), 67–90 (2020)

63. ChipSoft: HiX Homepage (2020). https://www.chipsoft.com/solutions/550/Solutions Accessed Accessed 18 Oct 2020

64. iMDsoft: MetaVision iMDsoft Homepage (2017). https://www.imd-soft.com/products/intensive-care Accessed Accessed 20 Aug 2020

65. Gaye A, I.J.e.a. Marcon Y: Datashield: taking the analysis to the data, not the data to the analysis. Int J Epidemiol. 43(6), 1929–1944 (2014)

66. Bizer, C., Heath, T., Berners-Lee, T.: Linked data - the story so far. Int. J. Semantic Web Inf. Syst. 5, 1–22 (2009)

67. Data, F.: FDP specification Homepage (2021). https://github.com/FAIRDataTeam/FAIRDataPoint-Spec Accessed Accessed 24 Aug 2020

68. Ontotext: GraphDB Homepage (2015). https://graphdb.ontotext.com/ Accessed Accessed 29 Jul 2021

69. Wilkinson, D.M.S.S.e.a. M.D.: Evaluating fair maturity through a scalable, automated, community-governed framework. Scientific Data 174(6) (2019)

70. Neo4j: Neo4j Graph Database Homepage (2019). https://neo4j.com/product/neo4j-graph-database/ Accessed Accessed 29 Jul 2021

71. Lysenko, R.I.A.S.M.e.a. A.: Representing and querying disease networks using graph databases. BioData Mining 9(23) (2016)

72. Neo4j: neosemantics (n10s): Neo4j RDF & Semantics toolkit (2019). https://neo4j.com/labs/neosemantics/ Accessed Accessed 29 Jul 2021

